# Identification and Validation of a Lipid Metabolism-associated Gene Signature for Predicting Survival in Sepsis Patients

**DOI:** 10.1101/2025.08.10.25333392

**Authors:** Chengxin Xue, Xinxin Xu, Siyu He, Zihan Yu, Xianying Lei, Haoyu Hou, Zhiming You, Qike Li, Yuduo Pu, Tao Xu, Chengli Wen

## Abstract

**Background:** Sepsis is a life-threatening condition associated with high mortality rates, claiming millions of lives globally each year. To improve prognostic prediction in sepsis, this study aimed to establish a lipid metabolism-associated gene signature for risk stratification and immune function evaluation.

**Methods:** **U**sing the sepsis dataset GSE65682 (Gene Expression Omnibus, GEO), lipid metabolism-associated genes were identified via GeneCards and intersection analysis. Hub genes selection integrated Univariate Cox, Least Absolute Shrinkage and Selection Operator (LASSO), and Multivariate Cox regression. Patients were stratified into high/low-risk groups by median risk scores. Prognostic performance was validated by Kaplan-Meier analysis and Receiver Operating Characteristic curves (ROC). Immune heterogeneity was analyzed using single-sample Gene Set Enrichment Analysis (ssGSEA), CIBERSORT, and correlation networks.

**Results:** A 9-gene prognostic signature (Aryl Hydrocarbon Receptor Repressor, AHRR; Ceroid-Lipofuscinosis, Neuronal 8, CLN8; Fatty Acid Synthase, FASN; Lanosterol Synthase, LSS; Mediator Complex Subunit 29, MED29; Platelet-Activating Factor Acetylhydrolase IB Subunit Alpha, PAFAH1B1; Phosphatidylinositol-4-Phosphate 5-Kinase Type 1 Gamma, PIP5K1C; Tribbles Pseudokinase 3, TRIB3; UDP-Glucose Ceramide Glucosyltransferase, UGCG) demonstrated robust predictive value. High-risk patients showed poor survival (KM, p=6.75 × e⁻⁸;ROC AUC: 0.951) and enriched Chemokine Receptor (CCR) and parainflammation, while low-risk individuals exhibited elevated higher infiltration levels of Tumor-infiltrating lymphocytes(TIL), type_II_IFN_Response, Treg, and macrophages . Immune network analyses revealed coordinated interactions: activated NK cells synergized with M1 macrophages (r=0.44) but antagonized resting NK cells (r=-0.62). Immune checkpoints CD86/TNFSF4 were upregulated in low-risk patients, contrasting with CD200R1 suppression.

**Conclusion:** This study successfully established a lipid metabolism-derived gene signature that provides a clinically actionable tool for prognostic stratification in sepsis patients, helping personalized clinical decision-making and facilitating early interventions.

## 1. Introduction

Sepsis, a life-threatening systemic inflammatory response to infection that often leads to multiple organ dysfunction syndrome (MODS), remains a global health challenge(Bai, Gao, Yan, & Zhao, 2024; Bauer et al., 2020; Singer et al., 2016). Globally, sepsis affects approximately 48.9 million people annually, causing 11 million deaths, while the U.S. healthcare system incurs economic costs nearing USD 38 billion each year due to this condition(Kumar, Balraj, Kempegowda, & Prashant, 2024).Traditional biomarkers exhibit insufficient specificity(He, Yue, Dong, Wang, & Cheng, 2024; Wolf, Wimalawansa, & Razzaque, 2019), highlighting the urgent need to develop novel prognostic biomarkers grounded in the pathophysiological mechanisms of sepsis.

Recent studies indicate that lipid metabolism dysregulation plays a pivotal regulatory role in the pathological progression of sepsis(Amunugama, Pike, & Ford, 2021). Research revealed that triggering receptor expressed on myeloid cells 2 (TREM2) binds to the Protein Tyrosine Phosphatase (PTPase)domain of phosphatase Src homology region 2 domain-containing phosphatase-1 (SHP1) to suppress Bruton’s tyrosine kinase (BTK) activation, thereby blocking fatty acid oxidation (FAO). This mechanism ultimately exacerbates inflammatory cytokine release and organ damage. This discovery first elucidates how TREM2 aggravates sepsis by inhibiting FAO through the "SHP1/BTK axis" (Ming et al., 2024). Notably, a proteomics study further confirmed that the lipoprotein metabolism-associated module (MEturquoise) in sepsis patients’ serum shows a significant correlation with disease severity. This module involves critical pathways such as plasma lipoprotein particle remodeling and cholesterol biosynthesis (Liang et al., 2021). Clinical epidemiological studies demonstrate that low-density lipoprotein cholesterol (LDL-C) levels exhibit a significant positive correlation with long-term incidence rates of community-acquired sepsis (Guirgis et al., 2016). In addition, a study found that through integrated analysis of single-cell RNA sequencing and metabolomic data, the five genes Mitogen-Activated Protein Kinase 14(MAPK14), Epoxide Hydrolase 2(EPHX2), BMX Non-Receptor Tyrosine Kinase(BMX), Fc Fragment Of IgE Receptor Ia(FCER1A), and Platelet Activating Factor Acetylhydrolase 2(PAFAH2) serve as central regulators of lipid metabolism dysregulation in sepsis(She et al., 2023). Intriguingly, lipoprotein family members display dual regulatory properties in sepsis: oxidized low-density lipoprotein exacerbates inflammatory responses through Toll-like receptor 4(TLR4) pathway activation (ZHA Qing et al., 2017), while enterically derived high-density lipoprotein (HDL)-associated enzyme complexes exert protective effects by neutralizing endotoxins (Han et al., 2021). Although the studies cited above demonstrate strong associations between lipid metabolism disorders and sepsis prognosis, systematic investigation into the prognostic value of lipid metabolism-associated genes (LMAGs) remains lacking. To enhance the prognostic assessment of sepsis, this study seeks to develop a lipid metabolism-associated gene signature for risk stratification, thereby establishing a theoretical foundation for evaluating the predictive value of lipid metabolism-related genes in determining clinical outcomes of sepsis patients.

## 2. Materials and Methods

### 2.1 Data Collection

The RNA-sequencing (RNA-seq) data were obtained from the Gene Expression Omnibus (GEO) database (https://www.ncbi.nlm.nih.gov/geo). We selected the GSE65682 dataset(Scicluna et al., 2018) for analysis, which originally included samples from sepsis patients. To ensure data quality, samples with missing diagnostic outcomes, incomplete 28-day survival records, and those classified as non-hospital-acquired pneumonia (non-HAP) or undiagnosed/unspecified cases were excluded.

### 2.2 Identification of Lipid Metabolism-Associated Genes

Lipid metabolism-associated genes were retrieved from the "Lipid Metabolism" gene set in the GeneCards database (https://www.genecards.org). These genes were intersected with the pre-filtered GSE65682 dataset for subsequent functional analysis.

### 2.3 Development and Validation of a Sepsis Prognostic Risk Model

This study developed a prognostic gene signature for sepsis using transcriptomic data from a sepsis cohort, Patients were randomly assigned to the training and validation sets in a 1:1 ratio, ensuring balanced sample sizes between the two groups. Prognostic gene signature involved three-stage regression analysis. Initially, univariate cox regression analysis (significance threshold P<0.05) identified genes significantly associated with 28-day survival. The least absolute shrinkage and selection operator (LASSO) Cox regression was then applied for variable compression: LASSO utilized an L1-penalty term to shrink regression coefficients, with the tuning parameter λ governing the degree of shrinkage (higher λ values enforce stronger contraction of coefficients toward zero). During this process, coefficients of non-critical genes were reduced to zero and excluded from the model, while retained genes formed a core subset. The optimal λ (lambda.min) was selected through 10-fold cross-validation by minimizing the partial likelihood deviance, effectively balancing model parsimony and predictive performance. Subsequently, a multivariate cox regression model was constructed using LASSO-selected genes, and stepwise regression (bidirectional optimization via the akaike information criterion, AIC) identified independent prognostic genes. A risk score formula was established as follows: Risk Score = Σ (β × Z-score normalized expression), where β represents the regression coefficients derived from the multivariate model. Patients were stratified into high- and low-risk groups based on the median risk score. Model performance was evaluated through Kaplan-Meier analysis (median risk cutoff) and receiver operating characteristic curve (ROC) analysis. Finally, a forest plot illustrated the hazard ratios (HR) and 95% confidence intervals (CIs)of key genes, highlighting their independent prognostic contributions.

### 2.4 Divergent Immune Landscapes in Prognostically Segregated Sepsis Cohorts

#### 2.4.1 Heterogeneity of Immune Microenvironments Across Risk-Stratified Sepsis Subgroups

The immune microenvironment features of sepsis risk-stratified subgroups were systematically analyzed via single-sample gene set enrichment analysis (ssGSEA). The normalized ssGSEA enrichment score matrix was processed by grouping samples according to risk stratification (low/high). To eliminate non-informative variables, immune features with zero standard deviation across rows were filtered out. Row-normalized Z-scores were computed to standardize the data, and a heatmap was generated using the pheatmap R package. A blue-white-red gradient color scheme (colorRampPalette) was employed to visualize enrichment levels. To preserve the original risk subgroup order, column clustering was disabled (cluster_cols=F), and sample annotation bars were incorporated to demarcate subgroup classifications. Enrichment scores for immune cell types and pathways were visualized, with statistical significance defined as a two-tailed p-value <0.05.

#### 2.4.2 Hub Gene–Immune Feature Correlation Network Analysis

Based on gene expression profiles and immune cell infiltration data, this section employed spearman’s rank correlation analysis to investigate the intrinsic relationships between key genes and the immune microenvironment. Standardized gene expression matrices, hub genes, and immune cell abundance matrices calculated by the ssGSEA algorithm were integrated, followed by sample intersection matching to ensure data consistency. A dual-loop iteration was performed to systematically analyze each hub gene against 28 immune cell subtypes. The cor.test function was used to compute Spearman correlation coefficients and significance p-values, with statistical significance determined using a two-tailed hypothesis framework. Significance thresholds were annotated as ***p<0.001, **p<0.01, *p<0.05. Finally, a heatmap was generated via the ggplot2 package to visualize the network relationships, with red-white-blue gradients representing correlation strength. The axes displayed hub genes and immune features, while the legend integrated statistical significance markers and correlation scales, elucidating regulatory associations between key genes and the immune microenvironment. A modular heatmap was generated via the ggplot2 package to visualize correlation patterns across stratified gene–immune interactions.

#### 2.4.3 CIBERSORT-Based Immune Cell Interaction Network

This section constructed an immune cell interaction network based on CIBERSORT-derived immune cell infiltration proportion data. Following standardized preprocessing of raw data, zero-variance features were identified and removed by calculating standard deviations across immune cell subsets to eliminate non-informative variables. Missing values were addressed using the listwise deletion method to ensure data integrity. The cleaned standardized matrix was analyzed through Pearson correlation coefficients to evaluate linear associations among 22 immune cell subsets, with hierarchical clustering algorithms (via the corrplot package in R) optimizing the correlation matrix structure. Visualization employed a half-matrix heatmap to delineate interaction patterns, utilizing a gradient color scale to illustrate positive-negative correlation intensities, while incorporating correlation coefficients within the heatmap to enhance quantitative interpretation.

#### 2.4.4 Differential Analysis of Immune Cell Infiltration Scores Based on CIBERSORT

The differential analysis of immune cell infiltration scores based on CIBERSORT was performed using R for data processing and visualization. First, a matrix of infiltration scores for 22 immune cell subtypes was generated using the CIBERSORT algorithm, retaining samples with a P-value < 0.05. To ensure data uniqueness, sample IDs were standardized, and duplicate entries were removed using the avereps function. Processed immune infiltration data were integrated with clinical risk score data through sample matching, resulting in a combined dataset comprising high-risk and low-risk groups. For differential analysis, grouped boxplots were constructed using the ggpubr package. Data were restructured via the melt function, and ggboxplot visualized the distribution differences of immune cell subtypes between risk groups. Wilcoxon tests were applied to assess intergroup significance, with asterisks denoting statistically significant differences (***P < 0.001, **P < 0.01, *P < 0.05). In correlation analysis, spearman’s rank correlation coefficient was used to evaluate associations between risk scores and immune infiltration levels. For subtypes showing significant correlations (P < 0.05), scatterplots with trend lines were generated using ggplot2, and marginal density distributions from the ggMarginal package were overlaid to create bivariate joint distribution visualizations. Key dependencies included the limma, reshape2, and vioplot packages.

#### 2.4.5 ssGSEA-Based Differential Immune Pathway Activity Scores

The section evaluated immune pathway activity scores based on the ssGSEA algorithm by integrating genomic expression data with immune signature gene sets, systematically analyzing differences in immune pathway activity between distinct risk groups. A standardized dataset was constructed by calculating immune pathway activity score matrices for each sample from preprocessed gene expression profiles through transposition and renaming operations. Cross-validation methods were employed to align risk score data with immune score matrices, ensuring sample consistency and yielding an integrated dataset containing risk scores, risk groups, and corresponding immune pathway activity. For differential analysis, data reshaping techniques were applied to transform wide-format data into long-format structures suitable for visualization. Risk group factor variables were established to compare low-risk and high-risk groups. Non-parametric statistical analysis using the wilcoxon rank-sum test was performed, with boxplots generated via the ggpubr package to visualize immune pathway activity distributions between risk groups. Statistical significance was denoted using a three-tier asterisk system (***p<0.001, ** p<0.01, *p<0.05) to identify pathways with significant activity differences. The entire analytical workflow was implemented using bioinformatics-specific R packages (limma, reshape2), ensuring methodological reliability and result reproducibility.

#### 2.4.6 Immune Checkpoint Molecule Expression Profiling

By integrating immune checkpoint gene expression profiles with patient risk score data, this section systematically analyzed the expression differences of immune checkpoint molecules between high-risk and low-risk groups. The gene expression matrix of sepsis samples was first standardized, and duplicate samples were removed using transposed matrices (via the limma package’s avereps function), followed by log2(x+1) transformation to improve data distribution. Common samples were selected for subsequent analysis by matching the risk score file with the immune checkpoint gene list (containing key molecules such as programmed cell death protein 1, PD-1 and cytotoxic T-lymphocyte-associated protein 4, CTLA-4). Wilcoxon rank-sum test was applied to compare the expression differences of immune checkpoint genes between high- and low-risk groups, with a significance threshold of p < 0.05 to identify statistically significant immune regulatory molecules. The reshape2 package was used to convert wide-format data into long-format, constructing a structured dataset containing risk groups, gene names, and expression levels. Grouped boxplots were generated using the ggpubr package, with "#0066FF" and "#FF0000" distinguishing low/high-risk groups. Asterisks were annotated to indicate significance levels (***p < 0.001, **p< 0.01, *p < 0.05).

#### 2.4.7 Association Analysis Between Prognostic Risk Scores and Immune Cells

This section conducted a systematic association analysis between prognostic risk scores and immune cell infiltration levels in sepsis patients. First, the CIBERSORT algorithm was employed to calculate the immune cell infiltration abundance matrix for each sample. Immune cell data with statistical significance were filtered by applying a P-value threshold (P < 0.05), followed by standardized processing of sample identifiers to remove low-quality samples. Prognostic risk score data were then integrated, and an exact sample name matching process generated a unified dataset containing immune cell composition and risk stratification. To investigate associations between risk scores and specific immune cell subtypes, spearman’s rank correlation analysis was utilized for bivariate correlation testing. For immune cell types demonstrating statistically significant correlations (P < 0.05), two-dimensional scatterplots were constructed using the ggplot2 package to visualize the distribution of their abundance relative to risk scores, with linear regression trendlines superimposed to highlight directional relationships. To further elucidate data distribution characteristics, kernel density curves were added along the margins of the scatterplots via the ggExtra package, creating composite visualizations.

#### 2.4.8 Hub Gene-Immune Cell Correlation Lollipop Plot

This section constructed a lollipop plot to visualize associations between 9 hub genes and immune cell subtypes by analyzing their correlation profiles. Using R language, correlation coefficients and p-value data were imported and processed. To enhance visualization efficacy, conditional functions were employed to map correlation coefficients to color gradients and proportional point sizes. Data point dimensions were stratified into five tiers based on absolute p-values, with larger markers indicating stronger correlations to emphasize statistical significance. Axis ranges were optimized to ensure complete data representation. Connecting segments between the coordinate origin and correlation coefficient positions formed "lollipop stems", where segment length visually encoded both direction (positive/negative) and magnitude of correlations. Automatic color and size assignments followed predefined mapping protocols, with statistically significant results (p<0.05) explicitly labeled using red-colored p-value annotations.

## 3. Results

### 3.1 Data Collection

The original dataset (GSE65682) contained 802 samples categorized into four groups: hospital-acquired pneumonia (HAP, n=84), community-acquired pneumonia (CAP, n=108), non-hospital-acquired pneumonia (non-HAP, n=33), and undiagnosed unspecified cases (not available, NA, n=577). Following the removal of non-HAP, undiagnosed/unspecified cases, and samples with incomplete clinical data, the remaining cohort included 84 HAP and 108 CAP patients. After applying exclusion criteria, the final dataset comprised 183 sepsis patients., enabling focused investigation of pneumonia-associated sepsis outcomes.

### 3.2 Identification of Lipid Metabolism-Associated Genes

The intersection of the GeneCards lipid metabolism gene set with the genes in the GSE65682 dataset yielded 2,052 lipid metabolism-associated genes. This refined gene subset was subsequently used for further analysis.

### 3.3 Development and Validation of Sepsis Prognostic Risk Model

The 183 sepsis patients were randomly allocated into balanced training (n=92) and validation (n=91) cohorts. Through a triple screening strategy encompassing univariable cox proportional hazards regression analysis, LASSO regression analysis, and multivariable cox regression analysis (Fig 1), this study successfully constructed a clinical prognostic prediction gene signature for sepsis comprising nine LMAGs.

**Fig 1.**
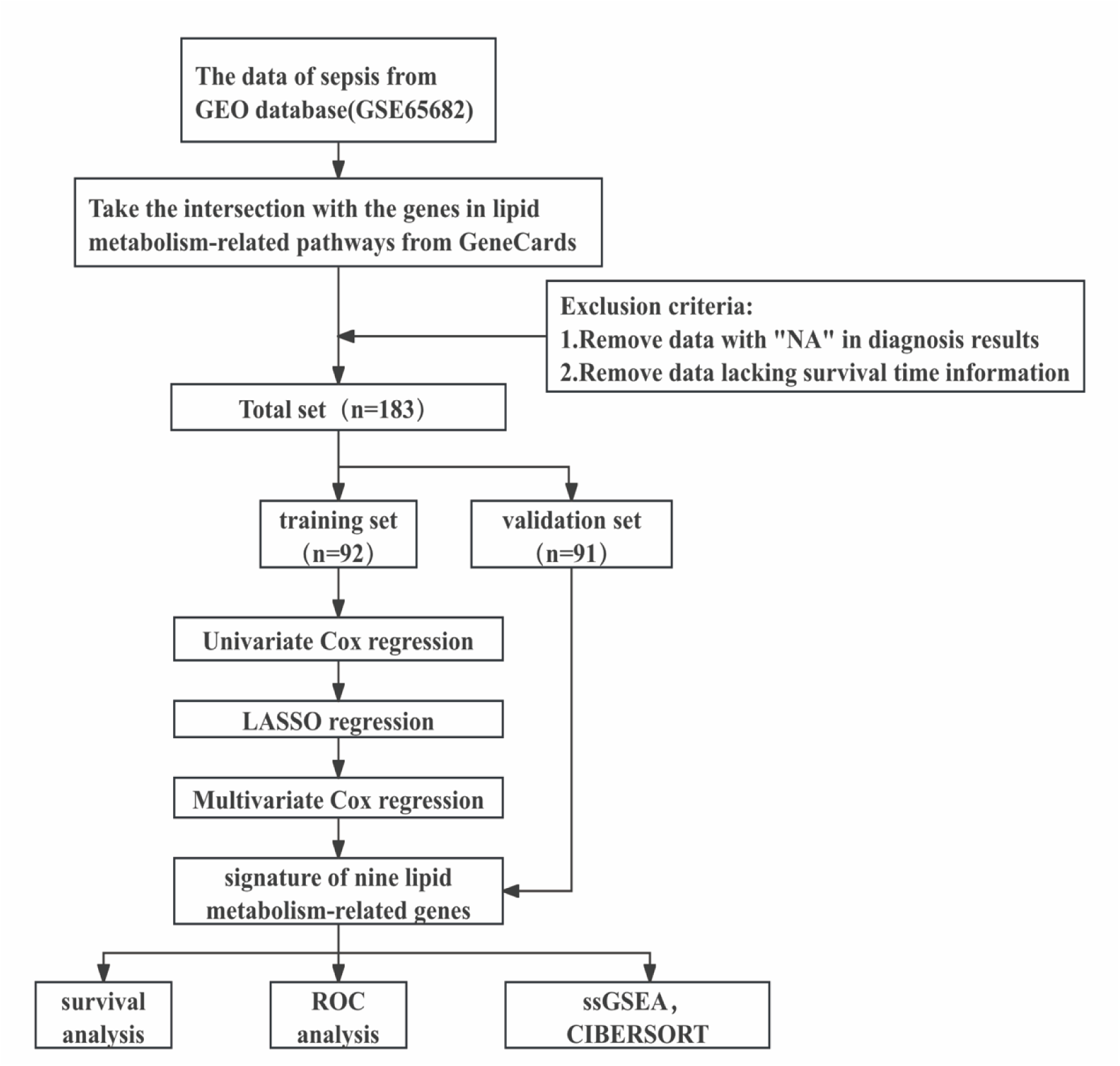
The flowchart of the research process.

Prognostic gene screening identified 940 survival-associated genes (P<0.05) through univariate cox analysis. Subsequent LASSO regression (Fig 2a and b) refined the candidates to 20 pivotal genes, including AHRR, ATP2A2, CLN8, COX19, CYP51A1, DECR1, FASN, GPCPD1, LIPA, LSS, MED29, NAAA, PAFAH1B1, PIP5K1C, PLIN2, PTGDS, ST3GAL5, TRIB3, UGCG.( Aryl Hydrocarbon Receptor Repressor ,AHRR; ATPase sarcoplasmic/endoplasmic reticulum Ca2+ transporting 2 ,ATP2A2;CLN8 transmembrane endoplasmic reticulum protein ,CLN8;Cytochrome c oxidase assembly protein COX19 ,COX19;Cytochrome P450 family 51 subfamily A member 1 ,CYP51A1;2,4-dienoyl-CoA reductase 1 ,DECR1;Fatty acid synthase ,FASN; Glycerophosphocholine phosphodiesterase 1 ,GPCPD1;Lipase A, lysosomal acid type ,LIPA; Lanosterol synthase ,LSS; Mediator complex subunit 29 ,MED29;N-acylethanolamine acid amidase ,NAAA; Platelet activating factor acetylhydrolase 1b regulatory subunit 1 ,PAFAH1B1;Phosphatidylinositol-4-phosphate 5-kinase type 1 gamma ,PIP5K1C;Perilipin 2 ,PLIN2;Prostaglandin D2 synthase ,PTGDS;ST3 beta-galactoside alpha-2,3-sialyltransferase 5 ,ST3GAL5;Tribbles pseudokinase 3 ,TRIB3; UDP-glucose ceramide glucosyltransferase ,UGCG)

**Fig 2.**
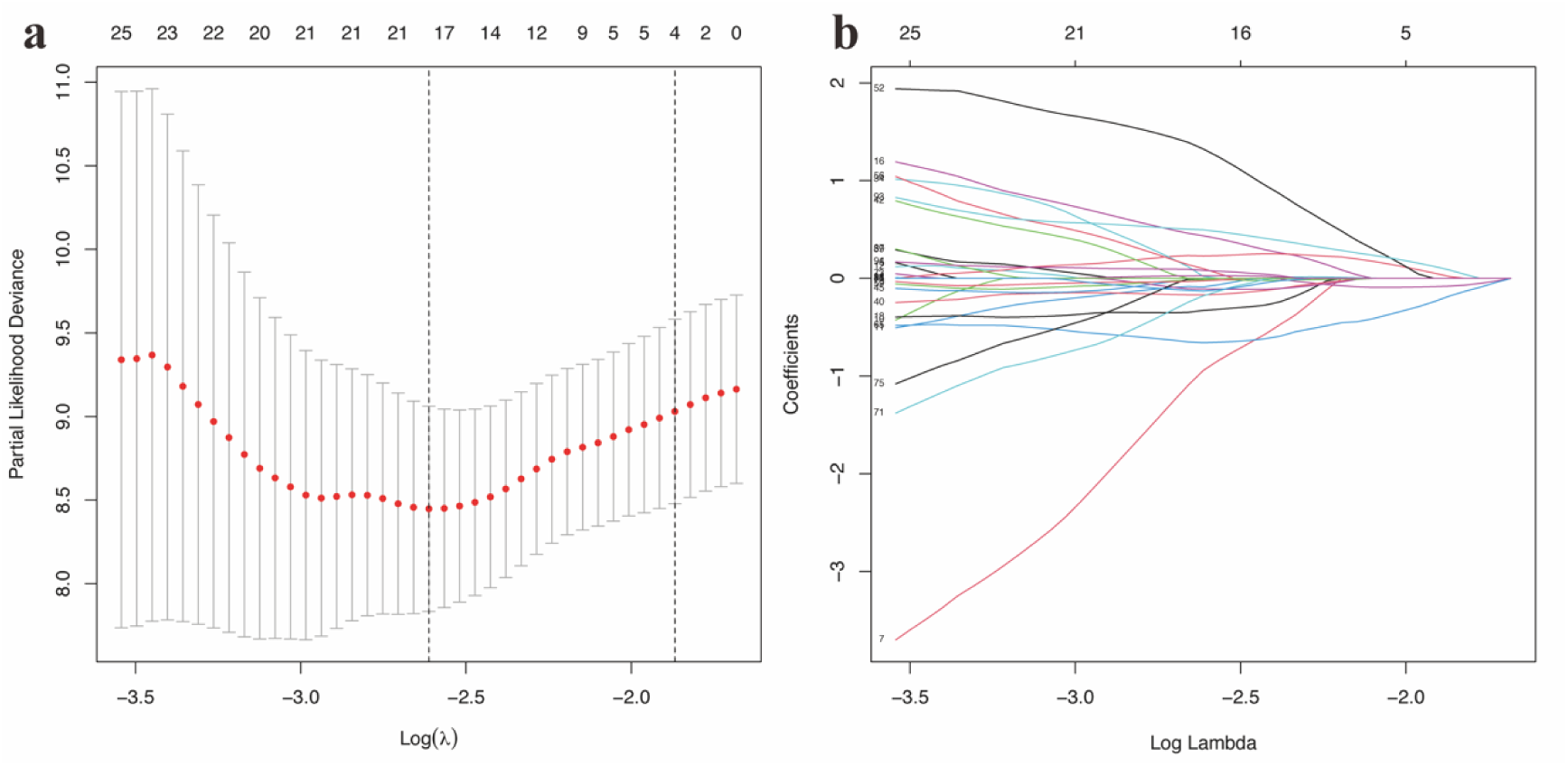
Construction of lipid metabolism-associated prognostic gene signature by LASSO regression analyses. a. The expression profiles of potential lipid metabolism-associated prognostic gene signature using LASSO coefficients. b. The penalty parameter (λ) of the LASSO model was chosen using ten cross-validation runs.

Multivariable Cox regression ultimately established a 9-gene prognostic signature comprising AHRR, CLN8, FASN, LSS, MED29, PAFAH1B1, PIP5K1C, TRIB3, and UGCG (Fig 3). (Aryl Hydrocarbon Receptor Repressor ,AHRR; Ceroid-Lipofuscinosis, Neuronal 8 ,CLN8;Fatty Acid Synthase ,FASN;Lanosterol Synthase ,LSS; Mediator Complex Subunit 29 ,MED29;Platelet-Activating Factor Acetylhydrolase IB Subunit Alpha ,PAFAH1B1;Phosphatidylinositol-4-Phosphate 5-Kinase Type 1 Gamma ,PIP5K1C;Tribbles Pseudokinase 3 ,TRIB3;UDP-Glucose Ceramide Glucosyltransferase ,UGCG)The mathematical formula for the risk score model is defined as: risk score = (-4.897 × AHRR) + (1.413 × CLN8) + (2.851 × FASN) + (3.449 × LSS) + (2.053 × MED29) + (-1.085 × PAFAH1B1) + (-2.750 × PIP5K1C) + (0.975 × TRIB3) + (0.669 × UGCG).This model demonstrated exceptional predictive performance, with an akaike information criterion (AIC) of 135.54 and a concordance index (C-index) of 0.93 (Fig 3).

**Fig 3.**
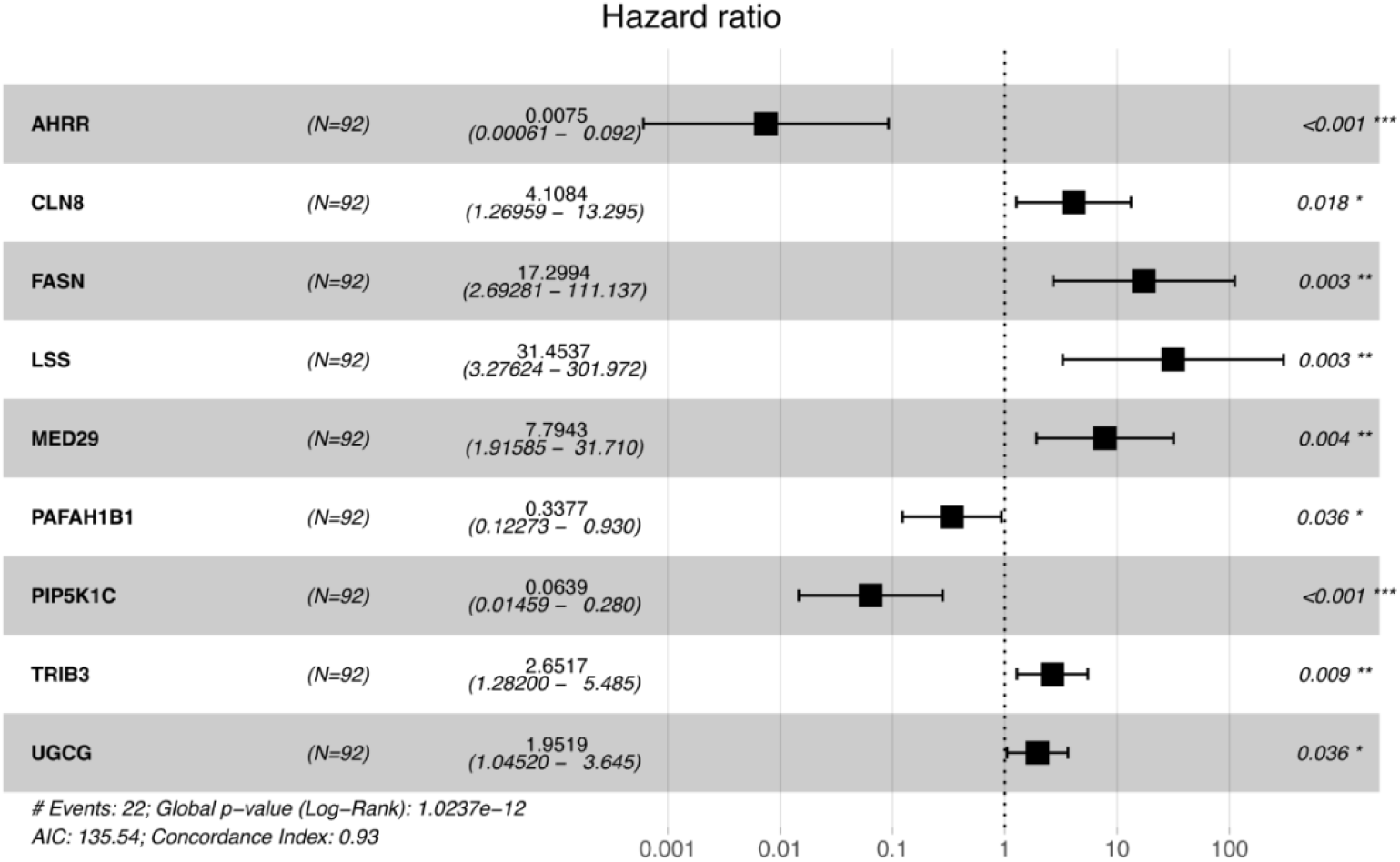
Multivariable cox proportional hazards regression analysis of hub genes associated with 28-day survival in sepsis. The forest plot demonstrates hazard ratios (HRs) with 95% confidence intervals, where statistical significance is annotated as ***P<0.001, **P<0.01, *P<0.05. Model robustness was evaluated using the akaike information criterion (AIC) to ensure optimal balance between predictive accuracy and parsimony.

In the training cohort, 92 sepsis patients were stratified into low-risk (n=46) and high-risk (n=46) groups based on the median risk score cutoff. Risk distribution visualization revealed distinct stratification characteristics between the two groups (Fig 4a–b). A differential expression heatmap confirmed significant heterogeneity in the expression patterns of the nine LMAGs across the groups (Fig 4c). Survival analysis indicated markedly superior 28-day survival rates in the low-risk group compared to the high-risk group (P = 6.75 × e⁻⁸, Fig 4d). ROC curve analysis further validated the model’s clinical utility, achieving an area under curve (AUC) of 0.951 for predicting 28-day survival (Fig 4e), underscoring its robust prognostic value.

**Fig 4.**
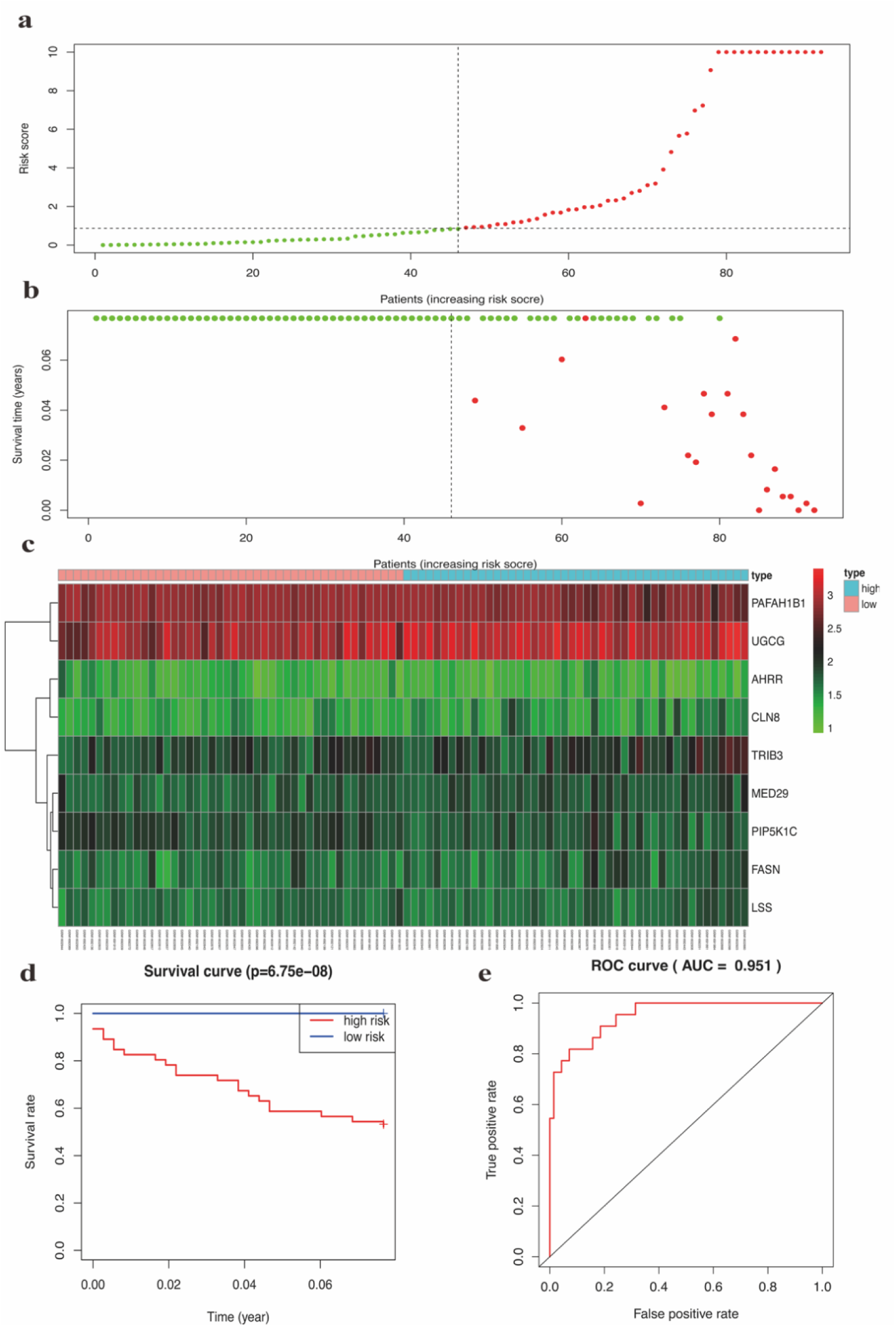
Prognostic performance of LMAG gene signature in the training set. a. Median risk score and distribution calculated by the LMAG-based model. b. Distribution of 28-d survival status in sepsis patients. c. Expression profiles of nine LMAGs between low- and high-risk subgroups. d. Kaplan-Meier analysis of 28-d survival stratified by LMAG-derived risk score. e. Prognostic accuracy of the risk score evaluated by AUC of the ROC curve.

In the validation cohort, sepsis patients were stratified into low-risk (n=61) and high-risk (n=30) groups using the same median risk score threshold as established in the training set (Fig 5a-b). The heatmap visualization of LMAG expression profiles across individual patients revealed distinct molecular signatures between the two risk groups (Fig 5c). For example, UGCG exhibited predominant upregulation in the high-risk sepsis subgroup. Subsequent Kaplan-Meier survival analysis demonstrated significantly higher 28-day survival probability in the low-risk group compared with their high-risk counterparts (P = 2.695 × e^-2^, Fig 5d). The risk score model exhibited predictive accuracy for 28-day mortality, achieving an area under the ROC curve of 0.655 (Fig 5e).

**Fig 5.**
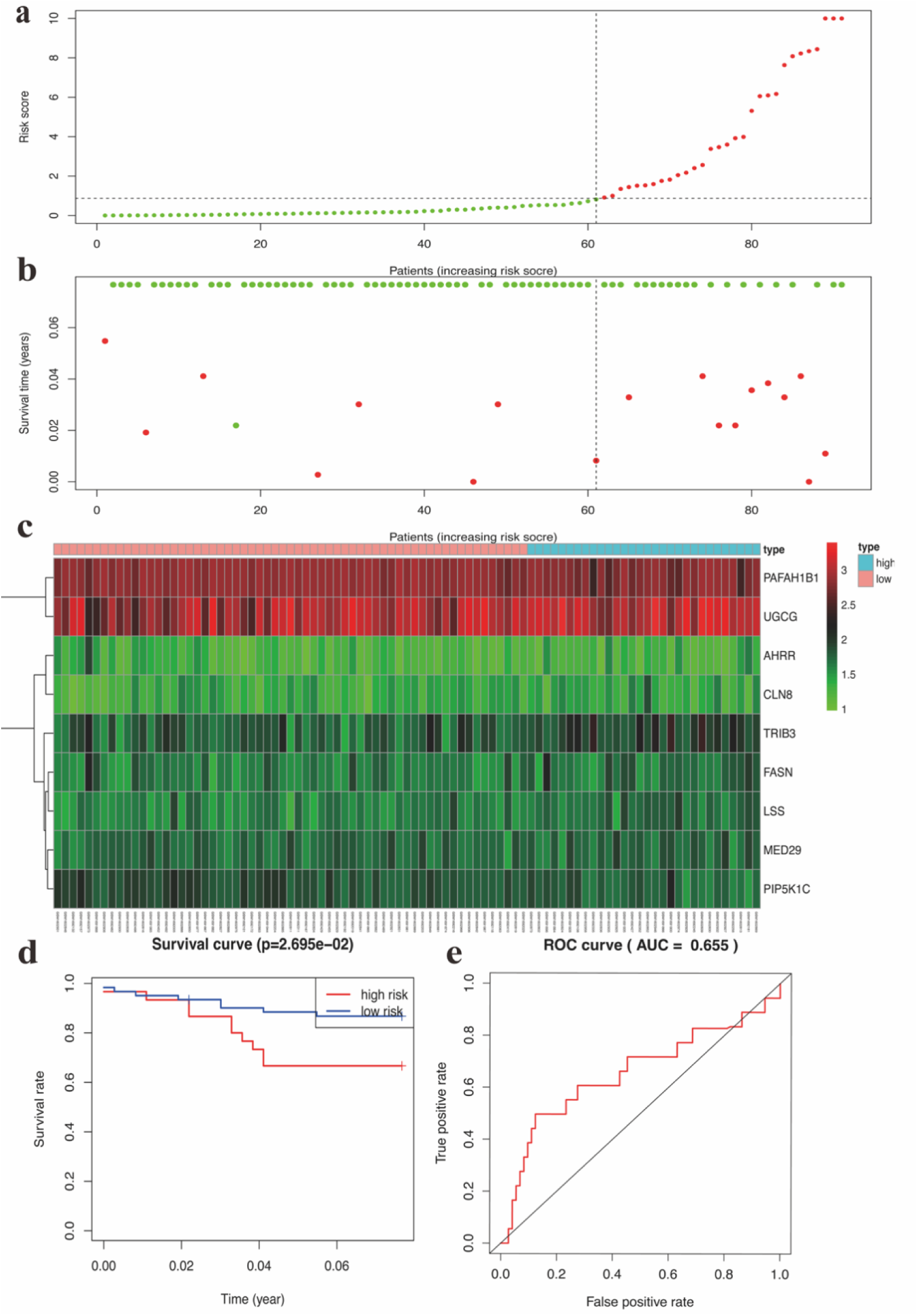
Prognostic performance of LMAG gene signature in the internal validation set. a. Median risk score and distribution calculated by the LMAG-based model. b. Distribution of 28-day survival status in sepsis patients. c. Expression profiles of nine LMAGs between low-risk and high-risk subgroups. d. Kaplan-Meier analysis of 28-day survival stratified by LMAG-derived risk score. e. Prognostic accuracy of the risk score evaluated by AUC of the ROC curve.

### 3.4 Divergent Immune Landscapes in Prognostically Segregated Sepsis Cohorts

#### 3.4.1 Heterogeneity of Immune Microenvironments Across Risk-Stratified Sepsis Subgroups

Firstly, based on the single-sample gene set enrichment analysis (ssGSEA) method, we systematically compared the differences in immune microenvironment characteristics between the low-risk and high-risk groups. The results revealed statistically significant differences (p<0.05) in eight immune-related features (Fig 6a): ① Th1 cells;② Tumor-infiltrating lymphocytes (TIL);③ Regulatory T cells (Treg);④Chemokine Receptor(CCR); ⑤ Neutrophils; ⑥ Para-inflammation; ⑦ Macrophages; ⑧Type II interferon response (Type_II_IFN_Response). Notably, the low-risk group exhibited higher infiltration levels of TIL, Type_II_IFN_Response, Treg, and macrophages, while the high-risk group showed significant enrichment of CCR and parainflammation.

**Fig 6.**
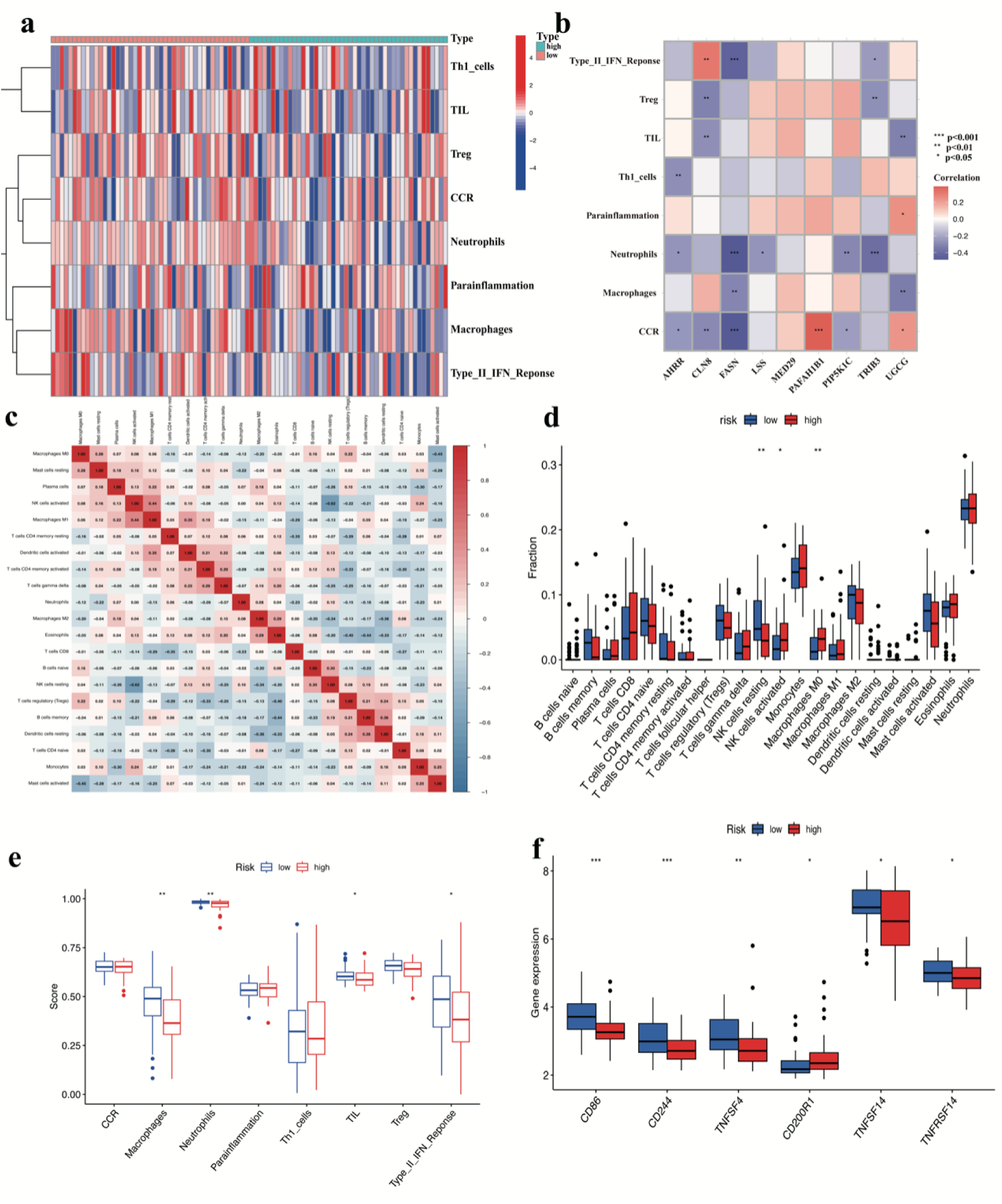
Heterogeneity of immune microenvironments across risk-stratified sepsis subgroups. a. ssGSEA-based characterization of immune microenvironment heterogeneity. b. Hub Gene–Immune Feature Correlation Network Analysis. c. CIBERSORT-Based Immune Cell Interaction Network. d. Differential analysis of immune cell infiltration scores based on CIBERSORT. e. ssGSEA-based Differential Immune Pathway Activity Scores. f. Immune Checkpoint Molecule Expression Profiling.

#### 3.4.2 Hub Gene–Immune Feature Correlation Network Analysis

Through comprehensive correlation network analysis of hub genes and immune features, this study revealed significant associations between 9 hub genes and 8 immune features (Fig 6b). The color gradient in each cell represents correlation coefficients, with red indicating positive correlations and blue denoting negative correlations. CLN8 showed a significant positive correlation with Type II interferon response (Type_II_IFN_Response) (p < 0.01), while FASN demonstrated a marked negative correlation with neutrophils (p < 0.001). Negative correlation patterns (blue clusters) predominated throughout the network architecture.

#### 3.4.3 Cibersort-based Immune Cell Interaction Network

The CIBERSORT algorithm-based deconvolution of immune cell interaction networks demonstrated significant functional synergy or antagonism among 22 immune cell subsets (Fig 6c). Notably, a robust positive correlation (Pearson’s r = 0.44, p < 0.05) was observed between activated natural killer NK cells (NK cells activated) and M1-polarized macrophages (Macrophages M1), suggesting their potential collaboration in pro-inflammatory microenvironments to amplify antisepsis or antimicrobial immune responses through cytokine crosstalk, such as the IFN-γ/IL-12 signaling axis.While, activated NK cells (NK cells activated) exhibited a strong functional antagonism (r = -0.62, p < 0.05) with resting NK cells (NK cells resting), a finding consistent with the biological paradigm of phenotypic switching during NK cell activation. This inverse relationship reflects the dynamic regulatory mechanism underlying the transition of NK cells from a quiescent, reservoir-like state to an effector cytotoxic phenotype.

#### 3.4.4 Differential Analysis of Immune Cell Infiltration Scores Based on CIBERSORT

CIBERSORT-based deconvolution analysis revealed significant differences in immune cell enrichment profiles between sepsis risk subgroups (Fig 6d). Comparative evaluation demonstrated that high-risk sepsis patients exhibited markedly elevated enrichment scores for activated NK cells ( P<0.05 ) and macrophages M0 ( P<0.01 ) compared to their low-risk counterparts. Conversely, resting NK cells showed significantly higher enrichment scores in the low-risk group (P<0.01).

#### 3.4.5 ssGSEA-Based Differential Immune Pathway Activity Scores

Through ssGSEA-based analysis of immune function enrichment scores, the results demonstrated that sepsis patients in the low-risk group exhibited significantly higher enrichment scores for neutrophil, macrophage, TIL, and Type II IFN Response compared to those in the high-risk group (Fig 6e).

#### 3.4.6 Immune Checkpoint Molecule Expression Profiling

Comparative analysis of immune checkpoint molecules revealed that sepsis patients in the low-risk group exhibited significantly elevated enrichment scores for cluster of differentiation 86(CD86)(P<0.001), cluster of differentiation 244 (CD244)(P<0.01), tumor necrosis factor superfamily member 4(TNFSF4)(P<0.01), tumor necrosis factor superfamily member 14(TNFSF14)(P<0.05) and tumor necrosis factor receptor superfamily member 14(TNFRSF14)(P<0.05) compared to the high-risk group, whereas the enrichment score of cluster of differentiation 200 receptor 1(CD200R1) (P<0.05) was markedly reduced (Fig 6f).

#### 3.4.7 Association Analysis Between Prognostic Risk Scores and Immune Cells

The scatter plot shows a significant correlation between the risk score and the level of immune cell infiltration. The abundance of M0 type macrophages (R = 0.33, P = 0.0014) and activated NK cells (R = 0.29, P = 0.0053) are positively correlated with the increase in the risk score (Fig 7a and c), suggesting that they may have potential significance as adverse prognostic factors. On the contrary, the proportion of activated mast cells (R = -0.24, P = 0.021) and the proportion of resting NK cells (R = -0.26, P = 0.012) are negatively correlated with the increase in the risk score (Fig 7 b and d).

**Fig 7.**
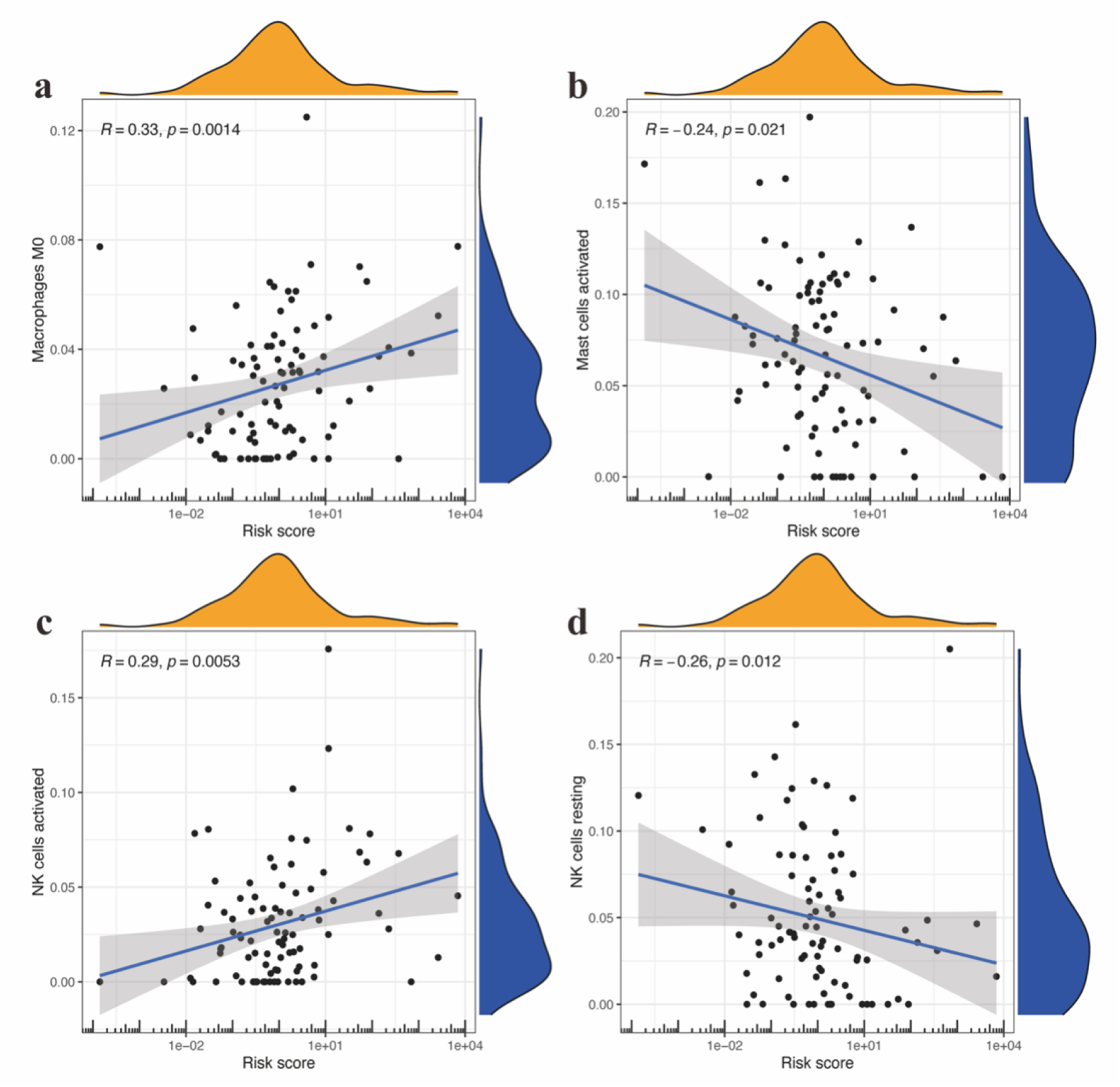
Immune cells content-risk score correlation plot. a. Macrophages M0 content-risk score correlation plot.b. Mast cells activated content-risk score correlation plot.c. NK cells activated content-risk score correlation plot.d. NK cells resting content-risk score correlation plot.

#### 3.4.8 Hub Gene-Immune Cell Correlation Lollipop Plot

Composite visualization revealed significant correlation features between 9 hub genes and 22 immune cell subtypes (Fig 8). When horizontally sorted by ascending |cor| values, a diagonal gradient pattern emerged across individual subplots (Fig 8a-i), with tight clustering of low-correlation features (|cor| < 0.2) near the origin gradually transitioning to more dispersed distributions in regions of higher |cor| values. For instance, the AHRR gene (Fig 8a) demonstrated strong positive correlations with Tregs(P=0.003), resting dendritic cells(P=0.003), and naive CD4+ T cells (P=0.02). Conversely, the inverse relationships with activated memory CD4+ T cells(P=0.039) and resting CD4+ T cells(P=0.039) may indicate antagonistic regulatory mechanisms, possibly reflecting differential gene regulatory networks during immune cell activation states.

**Fig 8.**
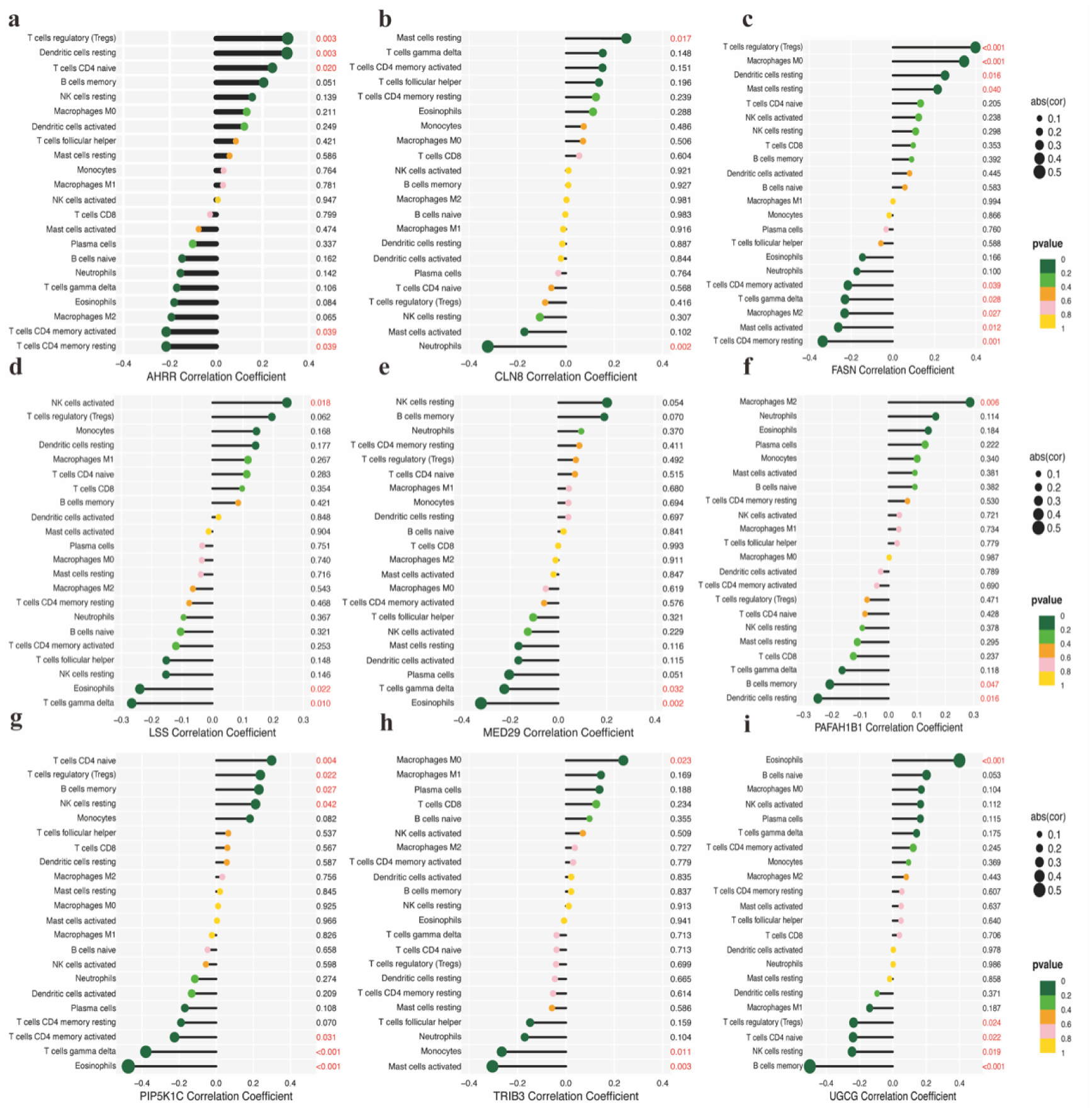
Hub gene-immune cell correlation lollipop plot. a. AHRR-immune cell correlation lollipop plot. b. CLN8-immune cell correlation lollipop plot. c. FASN-immune cell correlation lollipop plot. d. LSS-immune cell correlation lollipop plot. e. MED29-immune cell correlation lollipop plot. f. PAFAH1B1-immune cell correlation lollipop plot. g .PIP5K1C-immune cell correlation lollipop plot. h. TRIB3-immune cell correlation lollipop plot. i. UGCG-immune cell correlation lollipop plot. (Terminology clarification: cor refer to spearman’s rank correlation coefficient.)

## 4. Discussion

Sepsis is defined as a systemic syndrome characterized by multi-organ failure stemming from a dysregulated host response to pathogens(Markwart et al., 2020; Seymour et al., 2016). This pathophysiological process elevates mortality risks in intensive care patients(Abe et al., 2020), with affected patients representing approximately 30% of hospital admission cases (Rhee et al., 2019).

Through univariate cox regression, LASSO regression, and multivariate cox regression analyses, nine core genes (*AHRR, CLN8, FASN, LSS, MED29, PAFAH1B1, PIP5K1C, TRIB3, and UGCG*) were identified as critical factors in sepsis pathogenesis. Current evidence reveals their distinct mechanistic contributions. AHRR has been demonstrated to regulate intestinal epithelial lymphocyte (IEL) homeostasis through modulation of ferroptosis and oxidative stress responses(Panda et al., 2023). Its functional deficiency disrupts interleukin 10(IL-10) signaling pathways, exacerbating macrophage dysfunction during septic progression. FASN gene-mediated palmitoylation of myeloid differentiation primary response 88 (MYD88) serves as the critical molecular basis for Toll-like receptor (TLR) signaling activation(Kim et al., 2019). This pathway exacerbates sepsis progression by suppressing the chemotactic migration of neutrophils to infection sites. On the other hand, FASN regulates the palmitoylation modification of the STING protein by controlling malonyl-CoA levels(Kang et al., 2024). This newly discovered regulatory mechanism may offer a potential therapeutic target for ameliorating sepsis-associated hepatic injury. Low shear stress (LSS) has been mechanistically linked to endothelial barrier dysfunction through autophagic flux inhibition, characterized by reduced LC3B-II levels and p62 accumulation (Lupu et al., 2020). This pathophysiological process manifests as disrupted tight junction proteins (VE-cadherin, β-catenin) and cytoskeletal organization (F-actin disassembly), similarity to the glycocalyx shedding and intercellular junction breakdown observed in clinical sepsis(Lupu, Kinasewitz, & Dormer, 2020). The shear stress-endothelial dysfunction axis provides novel insights into sepsis-induced vascular permeability mechanisms. In contrast, PIP5K1C demonstrates stage-specific regulatory roles in sepsis progression through its modulation of phosphatidylinositol 4-phosphate (PI4P) metabolism and extracellular vesicle dynamics. LPS/TLR4 activation suppresses PIP5K1C expression via IFN-β signaling, triggering PI4P accumulation within multivesicular bodies and subsequent release of TNF-α-enriched pro-inflammatory vesicles. Sustained PIP5K1C inhibition creates a biphasic pathological trajectory: early-stage inflammation amplification through excessive extracellular vesicle (EV) secretion transitions to late-stage immune dysregulation as vesicle production becomes insufficient, mirroring clinical progression patterns observed in sepsis(Jin et al., 2023).Experimental evidence establishes TRIB3 as a direct prognostic factor through its suppression of cluster of differentiation 8-positive T lymphocyte cell(CD8+ T cell) infiltration. By repressing the signal transducer and activator of transcription 1 - C-X-C motif chemokine ligand 10 (STAT1-CXCL10 )axis, TRIB3 limits lymphocyte recruitment – a mechanism previously characterized in colorectal cancer immune evasion (Shang et al., 2022) that appears conserved in sepsis-associated immunosuppression. CLN8 (Settembre & Perera, 2024)and MED29 (Wu, Chen, Wu, & Fu, 2021)have demonstrated indirect sepsis associations through lysosomal dysfunction and extracellular signal-regulated kinase (ERK)/ nuclear factor kappa B (NF-κB) crosstalk, respectively. UGCG was consistently validated as a sepsis-related hub gene across cohorts (Ke et al., 2023; Li et al., 2023), While PAFAH1B1’s dual role in PAF hydrolysis reflects its context-dependent impacts on inflammation and thrombosis (Chang et al., 2015; Ward & Fattahi, 2019).

In the field of sepsis biomarker research, existing biomarkers generally suffer from limitations in timeliness or restricted predictive efficacy. C-reactive protein (CRP) and procalcitonin (PCT), the most widely used clinical biomarkers, exhibit prolonged detection windows and susceptibility to false positives in non-infectious inflammatory conditions, significantly diminishing their value in early diagnosis (Park et al., 2014). Identified biomarkers such as intestinal fatty acid binding protein (I-FABP) and D-lactate, while indicative of early intestinal injury, fail to effectively differentiate sepsis severity or predict clinical outcomes(X. Zhang, Liu, Wang, Yan, & Yang, 2019).Notably, high mobility group box 1 protein (HMGB1) demonstrates low clinical discriminative power with prognostic predictive AUC values ranging from 0.51 to 0.56 in severe sepsis patients(Karlsson et al., 2008). Although monocyte chemotactic protein-1 (MCP-1) and interleukin-6 (IL-6) possess certain prognostic value, their expression levels show significant temporal variations—MCP-1 demonstrates predictive utility only within the first three days of disease progression, while IL-6 exhibits substantial fluctuations in predictive stability across different timepoints (Barre et al., 2018).Compared to the aforementioned single biomarkers, the lipid metabolism-related gene signature established in this study demonstrates three distinct advantages through multi-gene synergistic mechanisms: ① Overcoming the temporal detection limitations of traditional biomarkers by enabling earlier sepsis risk identification; ② Integrating key regulatory nodes in lipid metabolism pathways to mitigate non-specific interference from single inflammatory factors; ③ Facilitating dynamic monitoring of metabolic reprogramming processes to achieve comprehensive disease course coverage from early warning to prognostic evaluation. This multidimensional prediction model based on metabolic network regulation provides a novel solution to overcome the clinical limitations of current sepsis biomarkers.

The findings of this study align with and mutually validate previous research: Firstly, the positive correlation between TIL scores and immune checkpoint gene expression, previously observed in solid tumors, was similarly identified in sepsis immune microenvironment analysis(H. Zhang et al., 2025). Our study revealed significantly elevated TIL infiltration levels in the low-risk group (p<0.05). At the immune regulation network level, single-cell sequencing studies have demonstrated the critical role of dendritic cells in regulating T cell differentiation through the interferon gamma receptor 2 ( IFNGR2 ) - signal transducer and activator of transcription 3 ( STAT3 ) signaling axis (Yao et al., 2022). The significant positive correlation between AHRR and dendritic cells (P=0.003) observed in our study suggests that lipid metabolism genes may participate in this regulatory network, potentially influencing T cell polarization through antigen-presenting cell modulation to improve sepsis outcomes. Notably, the role of lipid metabolic reprogramming in sepsis immune regulation was validated through multiple dimensions. The mechanism by which hub genes like MAPK14 influence prognosis through monocyte/macrophage differentiation (She et al., 2023) complements the lipid metabolism-related gene regulatory network identified in our study, collectively highlighting the pivotal position of the metabolism-immune axis in sepsis pathogenesis (Boomer et al., 2011). Clinically significant, the characteristic features of the low-risk group identified in our study - enhanced type II interferon response, dynamic Treg equilibrium, and macrophage activation - stand in stark contrast to the immunosuppressive phenotype observed in sepsis fatalities (characterized by reduced IFN-γ levels, hyperactivated Tregs, and abnormally elevated programmed death-ligand 1(PD-L1) expression in macrophages). This bidirectional evidence conclusively demonstrates the critical importance of maintaining immune homeostasis for prognosis improvement.

Although this study identified nine lipid metabolism-related core genes through multi-stage statistical methods and validated their prognostic predictive efficacy, certain limitations remain. Firstly, the conclusions are based on bioinformatics analysis and retrospective clinical data, lacking experimental validation of gene functions and mechanisms. Future studies plan to further explore these aspects through basic experiments: in vitro, CRISPR/Cas9 or siRNA technology will be employed to knockdown/overexpress target genes, evaluating their effects on lipid metabolism reprogramming, inflammatory cytokine release, and apoptosis pathways in sepsis- related cells such as macrophages and endothelial cells; in vivo, sepsis models using gene knockout mice will be constructed, with lipidomics analysis of hepatic/serum lipid metabolites combined with survival analysis and multi-organ injury biomarker detection to clarify the pathophysiological roles of target genes. Furthermore, the clinical applicability of the current predictive model requires validation through prospective studies. The next phase will involve conducting multicenter clinical cohort studies to dynamically monitor the expression profiles of these nine genes in peripheral blood mononuclear cells of patients. This will be correlated with lipoprotein metabolism indicators (e.g., HDL and LDL levels) to assess their association with sepsis severity and multi-organ failure development. Additionally, we will explore the combined predictive value of gene expression profiles with existing clinical scores (such as sequential organ failure assessment (SOFA) and acute physiology and chronic health evaluation II (APACHE II)). Long-term goals focus on developing rapid detection kits for key genes and screening small-molecule compounds or biologics that can regulate these targets, ultimately providing a theoretical foundation for precise sepsis subtyping and metabolic intervention therapies.

## 5. Conclusion

This study identified a lipid metabolism-associated gene signature through univariate cox analysis, LASSO regression analysis, and multivariate cox analysis, which demonstrated unique predictive value in the prognostic assessment of sepsis. Further mechanistic investigations revealed that sepsis progression is closely associated with dynamic changes in the immune microenvironment. These findings highlight the critical role of lipid metabolism-associated genes in sepsis outcomes. Furthermore, our study establishes a multi-gene predictive model with translational value and offers novel insights for individualized prognostic evaluation and precision therapeutic strategies.

## Data Availability

The RNA-sequencing (RNA-seq) data were obtained from the Gene Expression Omnibus (GEO) database (https://www.ncbi.nlm.nih.gov/geo). We selected the GSE65682 dataset(Scicluna et al., 2018) for analysis

## Abbreviations

AHRR: Aryl Hydrocarbon Receptor Repressor;
APACHE II: acute physiology and chronic health evaluation II;ATP2A2,ATPase sarcoplasmic/endoplasmic reticulum Ca2+ transporting 2;
AUC: area under the curve;
BMX: BMX Non-Receptor Tyrosine Kinase;
CCR: Chemokine Receptor;
CD200R1: cluster of differentiation 200 receptor 1;
CD244: cluster of differentiation 244;
CD8+ T cell: cluster of differentiation 8-positive T lymphocyte;
CD86: cluster of differentiation 86;
C-index: concordance index;
CIs: confidence intervals;
CLN8: Ceroid-Lipofuscinosis, Neuronal 8;
COX19: Cytochrome c oxidase assembly protein COX19 ;
CRP: C-reactive protein;
CTLA-4: cytotoxic T-lymphocyte-associated protein 4;
CXCL10: C-X-C motif chemokine ligand 10;
CYP51A1: Cytochrome P450 family 51 subfamily A member 1;
DECR1: 2,4-dienoyl-CoA reductase 1;
DEG: differentially expressed gene;
EPHA2: Epoxide Hydrolase 2;
ERK: extracellular signal- regulated kinase;
EV: extracellular vesicle;
FAO: fatty acid oxidation;
FASN: Fatty Acid Synthase;
FCER1A: Fc Fragment Of IgE Receptor Ia;
GEO: Gene Expression Omnibus;
GPCPD1: Glycerophosphocholine phosphodiesterase 1;
HAP: hospital- acquired pneumonia;
HDL: high-density lipoprotein;
HMGB1: high mobility group box 1 protein;
HR: hazard ratios;
IFNGR2: interferon gamma receptor 2;
I-FABP: intestinal fatty acid binding protein;
IL-10: interleukin 10;
LASSO: Least Absolute Shrinkage and Selection Operator;
LDL-C: low-density lipoprotein cholesterol;
LIPA: Lipase A, lysosomal acid type;
LMAGs: lipid metabolism-associated genes;
LSS: Lanosterol Synthase;
MAPK14: Mitogen-Activated Protein Kinase 14;
MCP-1: monocyte chemotactic protein-1;
MED29: Mediator Complex Subunit 29;
MODS: multiple organ dysfunction syndrome;
MYD88: myeloid differentiation primary response 88;
NA: not available;
NAAA: N-acylethanolamine acid amidase;
NF-κB: nuclear factor kappa B;
non-HAP: non-hospital-acquired pneumonia;
PAFAH1B1: Platelet Activating Factor Acetylhydrolase 1B Regulatory Subunit 1;
PAFAH2: Platelet Activating Factor Acetylhydrolase 2;
PD-1: programmed cell death protein 1;
PD-L1: programmed death- ligand 1;
PCT: procalcitonin;
PI4P: phosphatidylinositol 4-phosphate;
PIP5K1C: Phosphatidylinositol-4-Phosphate 5-Kinase Type 1 Gamma;
PLIN2: Perilipin 2 ;
PTGDS: Prostaglandin D2 synthase;
PTPase: Protein Tyrosine Phosphatase;
RNA-seq: RNA-sequencing;
ROC: receiver operating characteristic curves;
SOFA: sequential organ failure assessment;
ssGSEA: single-sample Gene Set Enrichment Analysis;
STAT1: signal transducer and activator of transcription 1;
STAT3: signal transducer and activator of transcription 3;
ST3GAL5: ST3 beta-galactoside alpha-2,3- sialyltransferase 5;
TIL: Tumor-infiltrating lymphocytes;
TLR: Toll-like receptor;
TLR4: Toll-like receptor 4;
TNFRSF14: tumor necrosis factor receptor superfamily member 14;
TNFSF14: tumor necrosis factor superfamily member 14;
TNFSF4: tumor necrosis factor superfamily member 4;
TRIB3: Tribbles Pseudokinase 3;
UGCG: UDP- Glucose Ceramide Glycosyltransferase.

## Ethics approval

Not applicable.

## Consent for publication

Not applicable.

## Availability of data and materials

The data come from GEO database.

## Conflicts of interest

The authors declare that they have no conflicts of interest.

## Funding

This study was fund by Southwest Medical University and Xuyong County People’ s Hospital (grant number:2023XYXNYD16), the Sichuan Science and Technology Program (grant number: 2022YFS0626), Luzhou Science and Technology Bureau(grant number: 2024SYF164), and Beijing Liver and Guts Charity Foundation(grant number: iGandanF-1082024-RGG140).

## Author contributions

**Chengxin Xue:** Writing – original draft, Visualization, Methodology, Formal analysis, and Data curation. **Xinxin Xu:** Visualization, Methodology, Formal analysis, and Data curation; **Zihan Yu, and Siyu He:** Methodology and Formal analyses**. Xianying Lei**: Conceptualization. **Haoyu Hou, Zhiming You, Qike Li, Yuduo Pu:** Methodology. **Tao Xu:** Conceptualization, Project administration, Funding. **Chengli Wen:** Conceptualization, Project administration, Supervision, Writing – review and editing, and Funding acquisition.

## Acknowledgements

The authors would like to thank the Information Center of Southwest Medical University for their help in this study.

## Notes

### Competing Interest Statement

The authors have declared no competing interest.

### Clinical Trial

Not applicable.

### Funding Statement

Yes

### Author Declarations

Ethics approval Not applicable.

